# DOPA pheomelanin is increased in nigral neuromelanin of Parkinson’s disease and it exacerbates alpha-synuclein neurotoxicity

**DOI:** 10.1101/2022.06.30.22277063

**Authors:** Waijiao Cai, Kazumasa Wakamatsu, Fabio A. Zucca, Kai Yang, Niyaz Mohamadzadehhonarvar, Pranay Srivastava, Gabriel Holly, Luigi Casella, Shosuke Ito, Luigi Zecca, Xiqun Chen

**Affiliations:** Department of Neurology, Massachusetts General Hospital, Harvard Medical School, Boston, USA; Department of Integrative Medicine, Huashan Hospital; Institutes of Integrative Medicine, Fudan University, Shanghai, China; Institute for Melanin Chemistry, Fujita Health University, Toyoake, Japan; Institute of Biomedical Technologies, National Research Council of Italy, Segrate (Milan), Italy; Department of Chemistry, University of Pavia, Pavia, Italy; Aligning Science Across Parkinson’s (ASAP) Collaborative Research Network, Chevy Chase, USA

**Keywords:** Parkinson’s disease, pheomelanin, eumelanin, DOPA, dopamine, alpha-synuclein, neuromelanin

## Abstract

**Objective:** Neuromelanin (NM) of the human substantia nigra (SN) has long been proposed as a key factor contributing to dopaminergic neuron vulnerability in Parkinson’s disease (PD). NM consists of pheomelanin and eumelanin moieties. Evidence supports that pheomelanin and eumelanin possess distinct chemical and biological characteristics. The present study aimed to investigate the relative composition and specific roles of pheomelanin and eumelanin moieties of NM in PD.

**Methods:** Pheomelanin and eumelanin components of NM in postmortem SN tissues from patients with PD were assessed by chemical degradation methods and compared with those from control subjects as well as patients with Alzheimer’s disease (AD). Additionally, synthetic pheomelanin and eumelanin were used to investigate their differential impacts on dopaminergic neuronal survival in a mouse model of PD overexpressing alpha-synuclein in the SN.

**Results:** We identified increased L-3,4-dihydroxyphenylalanine (DOPA) pheomelanin and increased ratios of dopamine (DA) pheomelanin markers to DA in PD SN compared to the controls. Eumelanins derived from both DOPA and DA were reduced in PD group. Melanin markers were unaltered in AD SN compared to the controls. Furthermore, we showed exacerbated dopaminergic neurodegeneration by synthetic DOPA pheomelanin and attenuated DA deficit by synthetic DOPA eumelanin in an alpha-synuclein mouse model of PD.

**Conclusion:** Our study provides insights into the different roles of pheomelanin and eumelanin moieties in PD pathophysiology. It forms a foundation for further investigations on pheomelanin and eumelanin individually as biomarkers and therapeutic targets for PD.

## 1. Introduction

In substantia nigra (SN) and locus coeruleus (LC) of the human brain there are catecholaminergic neurons which accumulate particular autolysosomal organelles. These organelles contain neuromelanin (NM) pigment, lipid, and proteins (1, 2). With aging a continuous accumulation of these NM-containing organelles occurs in neurons of different brain regions, albeit in a lower amount compared to SN and LC, which remain the most heavily pigmented regions of the aged brain (1, 3). Loss of pigmented dopaminergic neurons and consequently depigmentation of SN is a pathological hallmark of Parkinson’s disease (PD), a common neurodegenerative disorder manifested clinically by resting tremor, rigidity, bradykinesia, and gait instability (4). L-3,4-dihydroxyphenylalanine (L-DOPA or DOPA in this report) is the most commonly used symptomatic treatment for PD (5).

Unlike the well-characterized biosynthesis of melanin in the periphery (i.e., skin), which begins with the oxidation of tyrosine by tyrosinase to DOPA and then dopaquinone, followed by the addition of cysteine (Cys) to form cysteinyldopa (Cys-DOPA) and eventually pheomelanin or eumelanin when cysteine (Cys) is absent (6, 7), NM in SN is considered mainly a non-^1^enzymatic product from dopamine (DA) iron-mediated auto-oxidation (2, 6, 8). NM has a complex structure with the following components: melanin portion, lipids among which dolichols are the most abundant species, proteins and other constituents including iron and other metals (7, 9). The melanic moiety of NM pigment has been identified as composed of benzothiazine (BT), benzothiazole (BZ) and dihydroxyindole units (10), which means that melanic component of NM consists of pheomelanin and eumelanin moieties. Indeed, we have previously performed chemical analyses to elucidate the structure of NM in the SN (1, 10, 11), which suggested that the melanic part of NM pigment in the SN is mainly derived from DA and Cys in a molar ratio of 2:1 (11).

Our previous studies also indicated DOPA as a precursor for NM in brain regions, including SN, LC, putamen, premotor cortex, and cerebellum(1, 12). It was recently suggested that various catecholic metabolites are incorporated into NM synthesis from the SN and the LC. These compounds are metabolites of DA and norepinephrine formed by oxidative deamination by monoamine oxidase followed by reduction/oxidation (12-14). The involvement of enzymes in NM synthesis has also been proposed, including tyrosine hydroxylase (TH), which converts tyrosine to DOPA and is a marker for dopaminergic neurons. However, to date a neuronal specific enzymatic synthesis pathway of NM has not been unequivocally demonstrated (7). It was reported that overexpression of human tyrosinase in rat SN results in age-dependent production of peripheral type of melanin within dopaminergic neurons (15). However, evidence supports that tyrosinase is not present in human SN neurons (2, 16-18). NM interacts with several metals (1), quinones from DA oxidation, dopaminergic neurotoxins such as MPTP (1-methyl-4-phenyl-1,2,3,6-tetrahydropyridine), and other xenobiotics (19), indicating active roles of NM in aging and diseases, especially PD. However, the exact role of NM appears to be complex and can be neuroprotective or neurotoxic, depending on the specific cellular context (7).

Structurally, the casing model of mixed melanogenesis in the periphery (20, 21), previously applied to brain NM, has been reformulated based on more recent evidence (22). Protein/peptide fibrils constitute the core of NM particles on which quinone molecules produced by oxidative stress react, producing a melanic coat. The initial process resembles pheomelanin formation, as it involves mainly the exposed cysteine thiols of the fibrils, but also histidines and possibly lysines (22). Then, a chain of oxidation/condensation reactions of DA produce oligomers of unstructured eumelanic type (22). This process therefore differs from eumelanin formation in peripheral melanins and explains the lack of the characteristic structural feature of 3.5 Å typical of layers of DA eumelanin oligomers (23). The only structural element evidenced by X-ray powder analysis of NMs from all brain regions is the 4.7 Å motif of cross-β structure of the fibrillar protein core. For the scope of the present investigation, it will be sufficient to consider the surface NM melanic component as consisting of an inner layer of pheomelanic composition surrounded by unstructured eumelanin type oligomers.

Functionally, eumelanin and pheomelanin differ not only in color but also in their redox, metal chelating, and free radical scavenging properties (24). Eumelanin is an antioxidant, more stable, and photoprotective; pheomelanin is more prone to photodegradation and can act as a pro-oxidant by either reducing antioxidants or generating reactive oxygen species (24). The chemical architecture of NM together with the distinct properties of pheomelanin and eumelanin suggest possibly distinct pathophysiological roles of the two major components of NM in PD (25). The present study aimed to determine levels of NM and its pheomelanin and eumelanin components in postmortem SN tissues from patients with PD and compare them with healthy control subjects. For additional control and comparison, we included patients with Alzheimer’s disease (AD), another common, age-related neurodegenerative disease that is not generally viewed to affect nigral dopaminergic neurons (26). Furthermore, we investigated the impact of synthetic pheomelanin and eumelanin that are components of NM on dopaminergic neuronal survival in a mouse model of PD overexpressing alpha-synuclein (αSyn) in the SN (27).

## 2. Methods

### 2.1. Study design

The objective of the study was to test the overall hypothesis that pheomelanin and eumelanin are differentially associated with PD. Specifically, postmortem tissues were used to test the pre-specified hypotheses that PD SN contains higher pheomelanin than CON subjects with no neurological conditions. AD serves as an additional control. AD typically affects cortex and hippocampus and is pathologically characterized by amyloid plaques and neurofibrillary tangles (26), which is different from PD. The postmortem samples were requested from the Harvard Brain Tissue Resource Center. Sample sizes were determined by previous NM studies from the co-authors and others using postmortem brain tissues (1, 2, 21, 28, 29), the hypothesized effect size at 15-20% for the primary outcome measure (pheomelanin markers), and available samples with matching age, PMI, and race. Pheomelanin and eumelanin levels were determined using published chemical methods by established groups of the co-authors’, who were blind to the sample group information. Measurements were repeated 2-3 times, and the final values were the average from the repeats. Statistical analysis was conducted in consultation with the Harvard Catalyst Biostatistics Consulting Program and Dr. Jian Wang at The University of Texas MD Anderson Cancer Center. No data were excluded.

To test the pre-specified hypothesis that pheomelanin is neurotoxic and eumelanin is neuroprotective, an established αSyn mouse model of PD was used. Sample sizes were determined based on our previous studies in which significant differences in primary outcome measures (nigral dopaminergic cell counts and striatal DA content) were observed (27, 30). Melanin treatment groups were randomly assigned among all the animals preinjected with αSyn. Control animals received vehicle treatment. The endpoint was prospectively selected based on our previous studies using the same mouse model (27). Dopaminergic neurons and DA levels were determined by methods that were routinely performed by the authors. All outcome assessments and quantification were conducted by the investigators who were blind to the sample group information. No data were excluded.

### 2.2. Human postmortem SN samples

Human postmortem ventral midbrain blocks containing SN were provided by the Harvard Brain Tissue Resource Center through the National Institutes of Health (NIH) NeuroBioBank. The cohort includes PD, n = 12, age 70 - 74 years old, PMI 13.9 - 35.4 hours; AD, n = 11, age 65 - 74 years old, PMI 6.5 - 26.5 hours; and CON with no neurological disorders, n = 8, 68 - 73 years old, PMI 18.0 - 32.7 hours. All subjects were Caucasian. The neuropathological assessment confirmed PD or AD diagnosis. The SNpc was dissected from midbrain blocks, and tissues were carefully and mechanically ground without buffer. Aliquots were weighed and frozen at -80 °C for the determination of NM concentration. The remaining aliquots of samples were weighed and freeze-dried for chemical degradation analyses. All study protocols were approved by the institutional review board at the Massachusetts General Hospital and the institutions of the co-authors’.

### 2.3. Determination of NM concentration

Multiple aliquots of ∼5 mg obtained from homogenized SNpc wet tissues were processed to extract and measure NM concentration according to a previously published spectrophotometric method (31). The method was markedly improved to remove interfering tissue components (32). Three samples from PD group had limited amount of SNpc tissue; we thus prioritized determination of pheomelanin and eumelanin markers and did not measure NM in these 3 PD samples. For each of the 9 PD, 11 AD, and 8 CON samples, NM value was the average from 2-3 replicates. The final values of NM concentrations were expressed as µg/mg dry tissue.

### 2.4. Chemical degradation and determination of pheomelanin and eumelanin markers

Aliquots of the freeze-dry SN tissue (ca. 5-7 mg) prepared as previously described were used to determine pheomelanin and eumelanin moieties of NM using our well-established chemical oxidation and reduction methods followed by HPLC detection of markers for pheomelanin and eumelanin (28, 33, 34). Samples were homogenized in water with Ten-Broeck glass homogenizer at a concentration of 10 mg/mL (if samples were < 5 mg, we used 0.5 mL water) and 100 µL aliquots were subjected to the chemical reactions. Samples were oxidized with 1.5 % H_2_O_2_/K_2_CO_3_ (AHPO). After the termination of reaction, the mixtures were left for 20 hours at 25 °C to induce secondary production of PTCA, PDCA, and TTCA, the oxidative products from DOPA eumelanin, DA eumelanin, and DA pheomelanin, respectively, followed by HPLC analysis with UV detection (33). For the reductive reaction, samples were heated with 57 % HI in the presence of H_3_PO_2_ at 130 °C for 20 hours. Levels of 4-AHP, the degradative product of DOPA pheomelanin, and 4-AHPEA, the degradative product of DA pheomelanin were analyzed by HPLC-ECD (34-36). DA and DOPA were analyzed by HPLC-ECD under the same conditions as for 4-AHP and 4-AHPEA, respectively. Both oxidative and reductive reactions were performed on two separate occasions and the averages were reported. Contents of eumelanin and pheomelanin markers were calculated by the comparison with standard solution, and presented as ng/mg dry tissue (34). We also calculated all variables using wet tissue weight and found high correlations between dry tissue and wet tissue values.

### 2.5. Synthesis of DOPA pheomelanin and eumelanin

DOPA eumelanin and DOPA pheomelanin were prepared according to the methods described in detail elsewhere (37). The melanin powder was dried by lyophilization and equilibrated with moisture in a desiccator with a saturated CaCl_2_ solution.

### 2.6. Animals

Adult (6-7 months old) male C57BL/6 mice were purchased from the Jackson Laboratory (Bar Harbor, ME). Animals were maintained in home cages at a constant temperature with a 12-hour light/dark cycle and free access to food and water. The animals’ care was in accordance with institutional guidelines. All animal protocols were approved by the Massachusetts General Hospital Animal Care and Use Committee.

### 2.7. AAV human WT αSyn intranigral injection

pAAV-CBA-human αSyn WT-WPRE were infused at a volume of 2 µl and titer of 7.8 × 1012 into the left SN at the coordinates of AP +0.9 mm, ML +1.2 mm, and DV -4.3 mm relative to lambda (27).

### 2.8. Synthetic DOPA melanin injection

Synthetic DOPA pheomelanin (DOPA:Cys = 1:1) and DOPA eumelanin (DOPA:Cys = 1:0) were suspended in sterile PBS and sonicated overnight. Mice were randomly divided into 3 groups 6 weeks after AAV injection to received 0.5 µg/2 µl suspension of either DOPA pheomelanin or eumelanin or vehicle PBS infusion into the left SN at the coordinates of AP +0.9 mm, ML +1.2 mm, and DV -4.3 mm relative to lambda (27). All animals were sacrificed 4 weeks after synthetic melanin injection.

### 2.9. Measurement of striatal DA and DOPAC

The striatum was dissected and DA content in the striatum was determined by HPLC-ECD as previously described (27).

### 2.10. TH immunohistochemistry and stereological analysis of nigral dopaminergic neurons

Hinder brain was dissected, fixed, and cryoprotected. The brain was sectioned coronally at 30 μm, and a complete set of serial midbrain sections were collected. Sections were then immunostained for TH (Enzo Life Sciences, BML-SA497-0100 at 1:1000). Unbiased stereological counting was performed as previously described (27).

### 2.11. p-αSyn immunohistochemistry

An additional set of midbrain sections were processed and p-αSyn was stained using antibody against pSer129-αSyn (BioLegend Cat# 825701 at 1:500). Positively stained particles in the SNpc were counted using the optical fractionator method at 40× magnification (Olympus BX51 microscope and Olympus CAST stereology software) (27, 38).

### 2.12. Fluorescent staining for iba1

For iba1 staining, sections were processed and incubated with anti-iba1 (clone EPR16588, Abcam, ab178846 at 1:2000) overnight at 4ºC and goat anti-rabbit lgG-Alexafluor-568 (Invitrogen # A-11035, 1:1000) for 1 hour at 37 ºC. Images were captured, and iba1-positive cells in the SNpc were analyzed based on their morphology classifications of resting, reactive, and phagocytic microglia as previously reported (27, 39). Posterior (interaural 0.00/bregma - 3.80 mm), posterior central (interaural 0.28 mm/bregma -3.52 mm), anterior central (interaural 0.64 mm/bregma -3.16 mm), and anterior (interaural 0.88 mm/bregma -2.92 mm) midbrain sections from each mouse were analyzed and total counts were reported (40).

### 2.13. Statistical analysis

All values were presented as mean ± SEM. For results using human postmortem samples, statistical analysis was performed using ANOVA to initially compare differences among groups and Tukey test for pairwise comparisons. Analysis of covariance (ANCOVA) was used to compare outcome variables among groups adjusting for gender or other confounding factors as indicated in Results. Linear correlations between analytes were estimated by Spearman rank correlations. For results from animal experiments, differences among groups were analyzed using one-way or two-way ANOVA and Tukey post hoc test. P<0.05 was considered statistically significant.

## 3. Results

### 3.1. Characteristics of study subjects and materials

We obtained postmortem SN samples from 12 patients with PD and 8 controls (CON) with no neurological conditions. We also included analysis of postmortem SN tissues from the same tissue bank from 11 patients with AD. Diagnosis of PD or AD was confirmed pathologically. There were no statistical differences in age and postmortem interval (PMI) among PD, AD, and CON subjects (Table 1). All subjects were Caucasian.

**Table 1.** Characteristics of the study subjects and materials.

|  | n | Age (year) | Gender | PMI (hour) |
| --- | --- | --- | --- | --- |
| CON | 8 | 71.8±0.5 | m=6, f=2 | 23.5±1.9 |
| AD | 11 | 71.7±1.1 | m=6, f=5 | 21.1±1.7 |
| PD | 12 | 71.6±0.7 | m=4, f=8 | 23.0±2.2 |

### 3.2. Reduced NM and DA in postmortem PD SN, not in AD SN

We dissected SN pars compacta (SNpc) and determined DA and its precursor DOPA in the postmortem SN tissues by high performance liquid chromatography (HPLC) coupled with electrochemical detection (ECD). Consistent with the pathological diagnosis, HPLC revealed markedly lower DA in SN of PD as compared with CON (p=0.001, Figure 1A). DOPA levels in PD was reduced by 30%, not significantly different from CON, which likely reflects exogenous DOPA from PD medications (Figure 1B). DA level was still significantly lower in PD (p=0.001) when DOPA level was adjusted to control for the confounding effect of exogenous DOPA in PD group. Surprisingly, AD patients displayed significantly decreased DOPA (p=0.019) compared to CON. There were no significant differences in DA and DOPA levels between PD and AD (Figure 1A&B). The conversion rate from DOPA to DA expressed as DA/DOPA was lower in PD compared with CON (p=0.020). The rate was unaltered in AD compared with CON (Figure 1C). It is noteworthy that we calculated DOPA after hydroiodic acid (HI) hydrolysis, which affords total DOPA, including Cys-DOPA, DOPA conjugated with proteins, as well as free DOPA. NM was isolated and its level was measured by spectrophotometry. Consistent with previous studies, PD patients showed decreased NM concentration in SN as compared with CON (p=0.032, Figure 1D). No significant difference in NM concentration was observed between AD and CON groups. Compared with AD, PD showed borderline reduced concentration in SN (p=0.050) (Figure 1D). NM correlated with DA in CON (r=0.833, p=0.010). This correlation is consistent with the current knowledge that NM is derived mainly from DA under normal conditions. Of note, NM and DA were determined independently in different laboratories of the co-authors’. The NM and DA correlation was lost in PD and AD groups (Figure 1E). DOPA-adjusted NM was still significantly lower in PD group as compared with CON (p=0.013).

**Figure 1:**
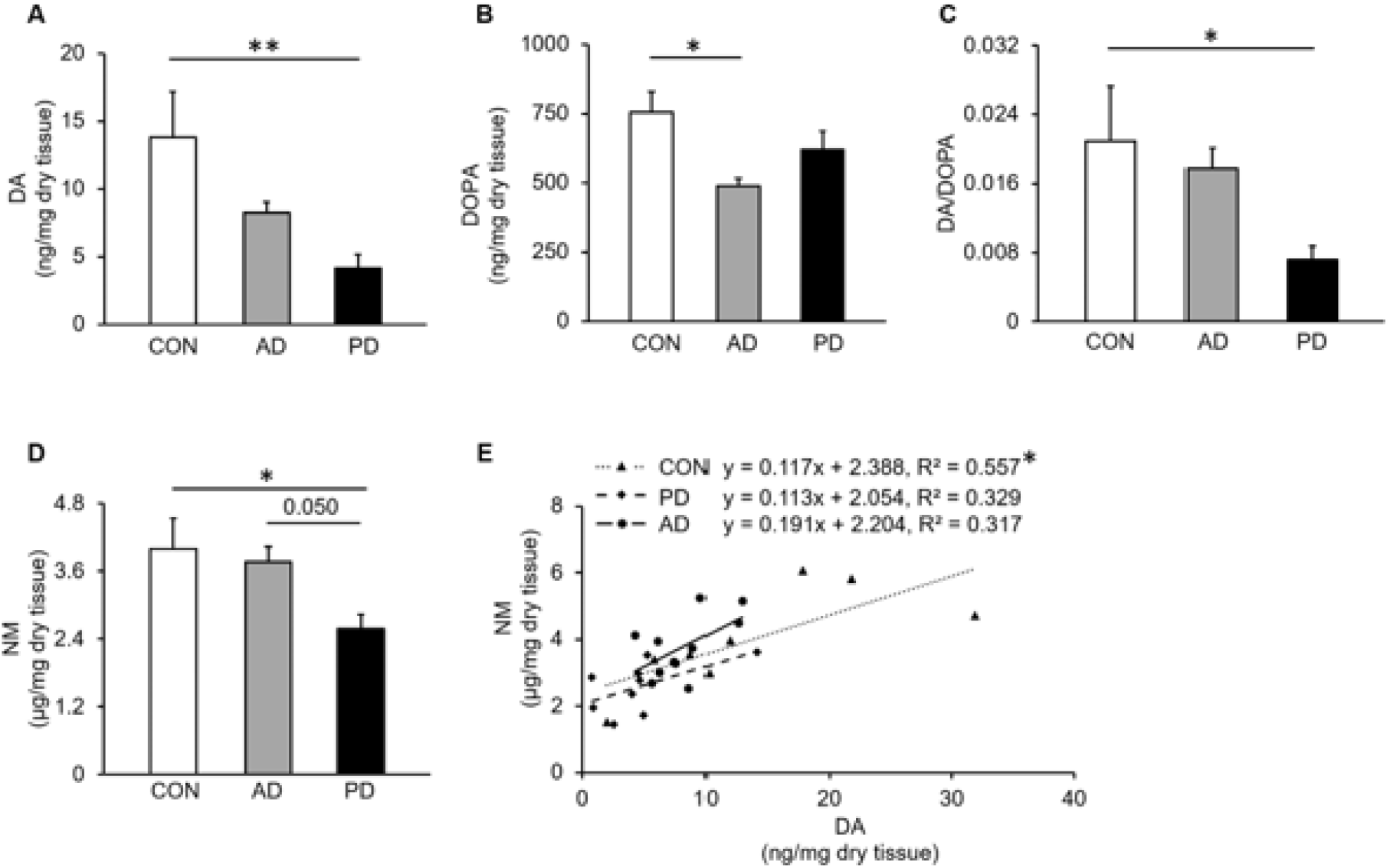
DA, DOPA, and NM in postmortem SN from PD, AD, or CON subjects. **(A)** DA levels determined by HPLC-ECD. **(B)** DOPA levels determined by HPLC-ECD. **(C)** DA to DOPA ratio. n=8, 11, and 12 for CON, AD, and PD, respectively (in **A-C**). **(D)** NM levels determined by spectrophotometry. **(E)** Correlation between NM and DA levels. n=8, 11, and 9 for CON, AD, and PD, respectively (in **D, E**). Values are mean ± SEM. *p<0.05, **p≤0.01, ANCOVA adjusting for gender followed by Tukey’s post hoc test or Spearman rank correlations.

### 3.3. Increased DOPA pheomelanin and reduced DOPA eumelanin and DA eumelanin in postmortem PD SN, not in AD SN

DOPA pheomelanin and DA pheomelanin in the SN were determined based on the formation of 4-amino-3-hydroxyphenylalanine (4-AHP) and 4-amino-3-hydroxyphenylethylamine (4-AHPEA), respectively, from HI reductive hydrolysis (28) (Supplementary Figure 1). HPLC-ECD analysis revealed a significantly higher 4-AHP level (p=0.025) and a trend for the higher rate of DOPA to DOPA pheomelanin conversion (4-AHP/DOPA ratio) in PD SN as compared to CON (Figure 2A). The difference in 4-AHP between PD and CON was stronger (p=0.003) when DOPA was adjusted to control for exogenous DOPA. There were no differences in 4-AHP and 4-AHP/DOPA ratio in AD group as compared to CON. Compared to AD, PD had a significantly higher 4-AHP level (p=0.015, Figure 2A). Level of 4-AHPEA, a BT unit marker for DA pheomelanin from HI reductive hydrolysis, was 59 % lower in PD SN as compared with CON (p=0.005, Figure 2B). However, PD patients showed a greater 4-AHPEA to DA ratio in SN than CON (p=0.042, Figure 2B). Level of 4-AHPEA in AD SN was not statistically different from CON, and AD group showed unchanged 4-AHPEA to DA ratio as compared to CON (Figure 2B). PD displayed no significant difference in 4-AHPEA from CON when DA was adjusted in the analysis, suggesting that the unadjusted difference may be explained by lower DA in PD despite the higher conversion rate as reflected by 4-AHPEA to DA ratio. Alkaline hydrogen peroxide (H_2_O_2_) oxidation (AHPO) was performed, and thiazole-2,3,5-tricarboxylic acid (TTCA), a BZ unit marker for DA pheomelanin, was determined by HPLC-UV detection (UVD). PD SN demonstrated a significantly lower level of TTCA as compared with CON (p=0.039, Figure 2C). However, a markedly higher TTCA/DA ratio was revealed in PD vs CON (p=0.012) and vs AD (p=0.008). TTCA and TTCA to DA ratio were not different in AD as compared with CON (Figure 2C). DOPA eumelanin marker pyrrole-2,3,5-tricarboxylic acid (PTCA) and DA eumelanin marker pyrrole-2,3-dicarboxylic acid (PDCA) from AHPO of the SN tissues were determined by HPLC-UVD. Reduced PTCA levels were revealed in PD SN as compared with CON (p=0.049). The borderline significance disappeared when DOPA was adjusted. PDCA was similarly lower in PD SN than CON with (p=0.016) or without adjusting for DA levels (p=0.010). PTCA and PDCA levels were not different in AD as compared with either CON or PD (Figure 2D&E). PTCA to DOPA and PDCA to DA ratios among all three groups were not significantly different (Figure 2D&E). As a result of changes in DOPA pheomelanin and DA pheomelanin, DOPA incorporation into pheomelanin as reflected by the ratio of 4-AHP/4-AHPEA was significantly higher in PD SN as compared with CON or AD (p=0.036 vs CON, p=0.005 vs AD), whereas DOPA eumelanin to DA eumelanin ratio PTCA/PDCA in PD SN was not different from CON and AD (Figure 2F). The ratio of pheomelanin/eumelanin is not only an indicator of NM composition but also melanin-related oxidative stress. A significantly greater 4-AHP/PTCA, which represents DOPA pheomelanin to DOPA eumelanin ratio, was revealed in PD SN as compared with CON or AD (p=0.011 vs CON, p=0.045 vs AD, Figure 2G). There was no difference in 4-AHP/PTCA between AD and CON. The 4-AHPEA to PDCA ratio, which reflects DA pheomelanin to DA eumelanin ratio from HI hydrolysis and AHPO, respectively, in PD SN and AD SN were not different from CON. Similarly, there was no difference among PD, AD, and CON groups in TTCA/PDCA, another DA pheomelanin to DA eumelanin ratio from AHPO (Figure 2G).

**Figure 2:**
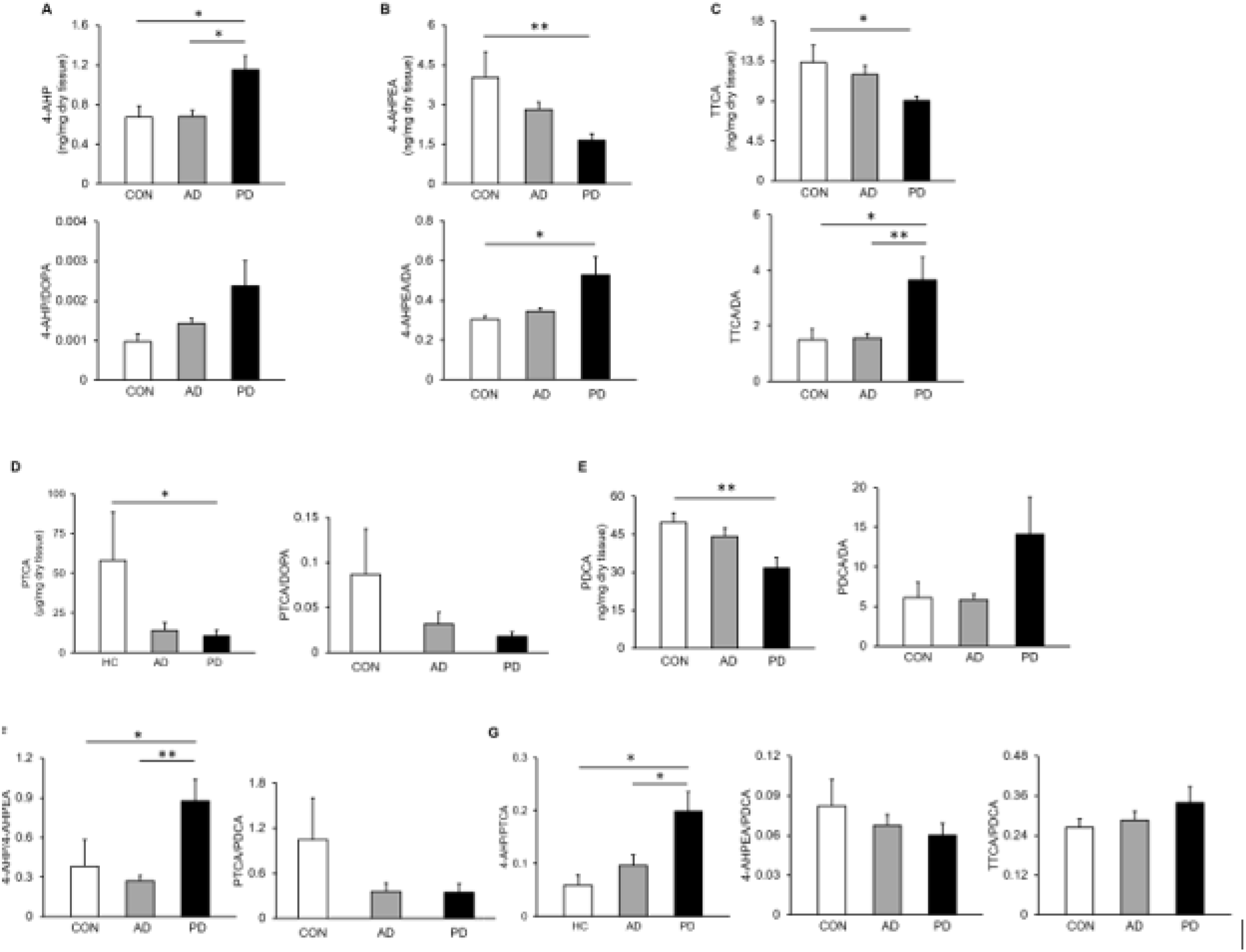
Pheomelanin and eumelanin in postmortem SN from PD, AD, or CON subjects. Postmortem SN tissues were subjected to HI hydrolysis or AHPO. **(A)** Levels of 4-AHP from HI hydrolysis, a marker for DOPA pheomelanin determined by HPLC-ECD, and 4-AHP to DOPA ratio. **(B)** Levels of 4-AHPEA from HI hydrolysis, a BT unit marker for DA pheomelanin determined by HPLC-ECD, and 4-AHPEA to DA ratio. **(C)** Levels of TTCA from AHPO, a BZ unit marker for DA pheomelanin determined by HPLC-UV, and TTCA to DA ratio. **(D)** Levels of PTCA from AHPO, a marker for DOPA eumelanin determined by HPLC-UV, and PTCA to DOPA ratio. **(E)** Levels of PDCA from AHPO, a marker for DA eumelanin determined by HPLC-UV, and PDCA to DA ratio. **(F)** 4-AHP to 4-AHPEA (DOPA pheomelanin to DA pheomelanin) ratio, and PTCA to PDCA (DOPA eumelanin to DA eumelanin) ratio. **(G)** 4-AHP to PTCA ratio (DOPA pheomelanin to DOPA eumelanin), 4-AHPEA to PDCA (DA pheomelanin to DA eumelanin) ratio, and TTCA to PDCA (DA pheomelanin to DA eumelanin) ratio. n=8, 11, and 12 for CON, AD, and PD, respectively. Values are mean ± SEM. *p<0.05, **p≤0.01, ANCOVA adjusting for gender followed by Tukey’s post hoc test.

### 3.4. Synthetic DOPA pheomelanin exacerbates dopaminergic neurotoxicity in an αSyn mouse model of PD

The above results from human study show that DOPA pheomelanin is more concentrated in NM of PD subjects than CON. To better understand roles of DOPA pheomelanin and eumelanin in neurodegeneration, we injected C57BL/6 mice with synthetic DOPA pheomelanin or DOPA eumelanin suspension or vehicle PBS into left SN 6 weeks after the mice received nigral injection of adeno-associated viruses (AAV) human wild type (WT) αSyn (27). Animals were sacrificed 10 weeks after the AAV injection. αSyn induced borderline non-significant DA deficit in the striatum (22%, p=0.051) and significant loss of TH-positive neurons in SN (29%, p=0.003) on the ipsilateral side as compared to the contralateral control side in vehicle-treated group (Figure 3A-B). The mild to moderate dopaminergic neurodegeneration was overall consistent with previous characterization of this model (27, 41). Synthetic DOPA pheomelanin administration resulted in a significant further decrease in the number of nigral TH-positive cells on the ipsilateral side compared with vehicle-treated group expressed either as absolute values (p=0.028) or percentage of the contralateral side (p=0.040). Synthetic DOPA eumelanin administration did not show a significant effect on dopaminergic neuron survival as compared with vehicle. Compared with pheomelanin-treated group, the number of TH-positive cells was higher on the ipsilateral side in the SN in eumelanin-treated animals (p=0.012, absolute values; p=0.010, percentage of the contralateral side) (Figure 3A).

**Figure 3:**
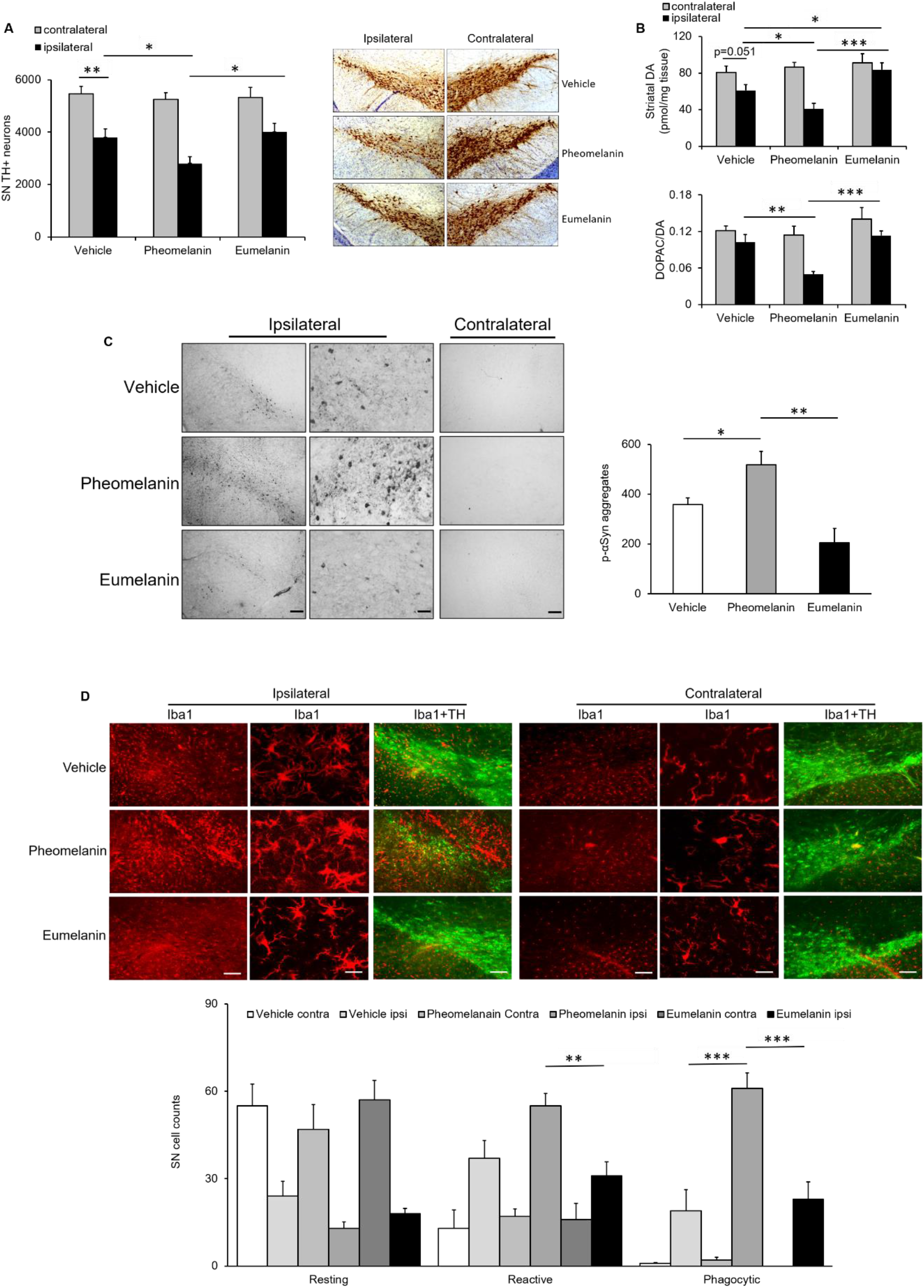
Synthetic DOPA pheomelanin exacerbates αSyn-induced dopaminergic neurotoxicity in AAV αSyn model of PD. Mice received AAV human WT αSyn injection into the left SN. Synthetic DOPA pheomelanin or DOPA eumelanin PBS suspension or vehicle PBS was injected into the ipsilateral SN 6 weeks after the AAV injection. Animals were sacrificed 10 weeks after AAV injection. **(A)** Stereological quantification of nigral TH-positive neurons (n=8, 9 and 10 for vehicle, DOPA pheomelanin, and DOPA eumelanin groups, respectively), and representative SN sections immunostained for TH. **(B)** Striatal DA determined by HPLC-ECD, and DOPAC to DA ratio. n=8, 9 and 10 for vehicle, DOPA pheomelanin, and DOPA eumelanin groups, respectively. **(C)** Immunoblotting for p-αSyn at Ser129 and quantitative analysis of p-αSyn aggregates in SNpc on the ipsilateral side. Scale bars = 20 µm, 8 µm (ipsilateral panels, from left to right), and 20 µm (contralateral panel). n=6/group. **(D)** Representative SN sections immunostained for iba1 and TH and morphology-based quantification of iba1-positive cell subtypes: resting (visible thin cytoplasm with long and thin processes), activated (dense and enlarged cell body with thick, short processes), and phagocytic (pseudo-amoeboid shape, big, dark cell body merging with processes) microglia. n=6/group. Scale bars = 50 µm, 10 µm, 50 µm (from left to right in the ipsilateral panels and contralateral panels). Values are mean ± SEM. *p<0.05, **p<0.01, ***p<0.001, two-way ANOVA (A, B, and D) or one-way ANOVA (C) followed by Tukey’s post hoc test.

Striatal DA content was lower on the ipsilateral side in DOPA pheomelanin-injected animals compared with vehicle-treated group (p=0.032, absolute values; p=0.035, percentage of contralateral side). In contrast, DA level on the ipsilateral side was not different from the contralateral side in DOPA eumelanin treated animals (Figure 3B) and higher compared with vehicle-treated animals (p=0.041) or DOPA pheomelanin-treated group (p<0.001, absolute values; p=0.002, percentage of the contralateral side) (Figure 3B). DA turnover rate (3,4-dihydroxyphenylacetic acid (DOPAC)/DA ratio) was not changed among all groups after normalizing to the contralateral side despite a significant reduction on the ipsilateral side in DOPA pheomelanin-treated animals compared to animals treated with either vehicle (p=0.001) or DOPA eumelanin (p<0.001) (Figure 3B). The exacerbated αSyn dopaminergic neurotoxicity following synthetic DOPA pheomelanin administration was accompanied by increased immunointensity for phosphorylated αSyn (p-αSyn) at Ser 129 on the ipsilateral side of the SN as compared with vehicle-treated mice (p=0.040). The accumulation of p-αSyn is central to the pathogenesis and progression of PD (42, 43). DOPA eumelanin did not alter p-αSyn aggregates in the ventral midbrain compared with vehicle group (p=0.053). Compared to pheomelanin, eumelanin-treated animals showed significantly reduced number of p-αSyn aggregates (p=0.007) (Figure 3C). Microglial responses to melanin treatment in the SN following αSyn overexpression were examined by analyzing the numbers of morphologically classified cells stained positive for ionized calcium-binding adapter molecule 1 (iba1). Although DOPA pheomelanin group did not show a difference in the number of reactive microglia as compared with ipsilateral vehicle control, reactive microglial cells were increased on the ipsilateral side in pheomelanin-treated group as compared with eumelanin group (p=0.007). In addition, DOPA pheomelanin induced a remarkable increase in the number of phagocytic microglia on the ipsilateral side as compared with vehicle control (p<0.001) and DOPA eumelanin group (p<0.001). The numbers of resting, reactive, and phagocytic microglia in eumelanin-treated animals were not statistically different from vehicle controls (Figure 3D).

## 4. Discussion

Pheomelanin and eumelanin possess distinct chemical and biological characteristics (44). Their metabolism and functions in the brain as the typical elements of NM are poorly understood, especially in the context of neurodegenerative disease. The present study identified in PD postmortem SN increased DOPA pheomelanin and increased conversion rates of DA to pheomelanin markers. Eumelanin derived from both DOPA and DA was decreased. In addition, we reported an increase in DOPA pheomelanin relative to DA pheomelanin in PD SN. In AD SN, we observed unaltered melanin markers despite reduced levels of DOPA. Furthermore, we showed exacerbated dopaminergic neurotoxicity by synthetic DOPA pheomelanin and attenuated DA deficit by synthetic DOPA eumelanin in the AAV αSyn mouse model of PD. The reduced DA melanin, both eumelanin and pheomelanin, in PD SN can be attributed to the substantially reduced DA levels at the advanced stage of the disease as a result of dopaminergic neuron degeneration, altered conversion from DOPA (since DA/DOPA was lower in PD SN), and altered DA metabolism (45, 46). This reduction is particularly evident for DA pheomelanin given the unaltered high correlations between Cys-DA pheomelanic moieties and DA in PD as compared to CON. However, the significantly higher ratios of TTCA to DA and 4-AHPEA to DA in PD SN still suggest enhanced DA oxidation and conversion to form prooxidant pheomelanic pigment. Accelerated DA oxidation in PD is supported by considerable literature evidence (15, 47, 48). Fornstedt et al. reported that, while SN DA was lower, the Cys-DA/DA ratio was substantially higher in SN in patients with degenerated SN dopaminergic neurons, and that the ratio seemed to be correlated with the degree of SN depigmentation and neurodegeneration (49). Spencer et al. later detected significant increases in cysteinyl adducts of DA in PD SN (50). Together with reduced DA eumelanin, these findings may indicate a shift of the NM production towards imbalanced pheomelanin and eumelanin and as results, oxidative stress despite the unchanged DA pheomelanin to eumelanin ratios.

Unlike DA, DOPA in PD SN was maintained at “normal” levels, likely by exogenous DOPA from DOPA therapies and reduced DOPA to DA conversion. The unaltered DOPA, however, is associated with a decrease in DOPA eumelanin and an increase in DOPA pheomelanin, suggesting further preferential production of pheomelanic over eumelanic DOPA pigment in PD SN in addition to the already favored pheomelanin production in NM synthesis based on casing model (20). DOPA treatment has been reported to lead to increased pheomelanogenesis and increased pheomelanin/eumelanin ratio in melanocytes *in vitro* (51). The abovementioned study from Spencer et al. also detected significant increases in cysteinyl adducts of DOPA in addition to cysteinyl adducts of DA in PD SN (50). Furthermore, the availability of Cys may be another important factor that facilitates the enhanced preference for pheomelanin production in PD (20). Cys normally is present in low concentrations in the brain, and pheomelanin biosynthesis could deplete Cys and related antioxidants (52, 53). However, increased Cys has been reported in neurodegenerative diseases (54), including PD likely due to the need to increase antioxidant protection.

Greater DOPA pheomelanin, together with reduced DA pheomelanin substantially increased DOPA pheomelanin accumulation in PD SN to ∼90 % of DA pheomelanin (4-AHP/4-AHPEA=0.88 in PD). Cause or effect of neurodegeneration in PD, or DOPA therapy-related, the higher levels of DOPA pheomelanin as well as reduced DOPA eumelanin and DA eumelanin in PD revealed dysregulated melanin production as well as redox homeostasis in PD SN (55). Although it remains to be determined how the changes in chemical composition in PD SN may affect the external structure of NM with its pheomelanic and eumelanic components on the surface, more pheomelanin and less eumelanin are ultimately associated with thinning protective surface (20, 21). Depletion of eumelanin will invariably lead to release of bound iron that, together with the exposure of the pheomelanin more redox reactive component will lead to more oxidative damage. The vicious circle between oxidative stress and neurodegenerative events in the SN likely plays a role in PD progression if not its initiation.

Our previous studies demonstrated that intranigral injection of NM isolated from the SN of human brain can cause nigral dopaminergic neuron loss in rats (56). It was suggested that NM accumulation that normally occurs in human SN dopaminergic neurons during aging (1, 4), eumelanin or pheomelanin notwithstanding, may increase expression and aggregation of the key PD protein αSyn in animal models as well as the human brain (2). Our demonstration of exacerbated dopaminergic neurotoxicity by synthetic DOPA pheomelanin in the AAV αSyn mouse model of PD provides direct evidence for a causal effect of pheomelanin and αSyn-induced dopaminergic neurodegeneration. The exacerbated αSyn dopaminergic neurotoxicity was associated with increased p-αSyn at Ser 129. The post-translational modification of αSyn can promote its aggregation and neurotoxicity (42, 43). In addition, DOPA pheomelanin exacerbated αSyn-induced microglial activation and phagocytosis. Our previous study indicated that human NM could activate microglia, which target neurons and their processes. Moreover, activated microglia release H_2_O_2_, NO and pro-inflammatory factors, which in the presence of massive release of bound iron, will cause further neurodegeneration with subsequent release of NM and microglia activation. This cascade produces a vicious cycle of neuroinflammation/neurodegeneration, which may contribute to progression of PD (56, 57). Despite unaltered microglial responses and restored DA deficit, DOPA eumelanin did not exert protection at the cell body level. The discrepancy between nigral dopaminergic neuron survival and striatal DA content may be due to enhanced DA production from existing dopaminergic neurons (58, 59) following eumelanin treatment. Alternatively, it may be possible that time courses for eumelanin impacts on dopaminergic neuron terminals and cell bodies are different. More investigations are needed to elucidate the mechanisms of pheomelanin and eumelanin actions.

The reduced DOPA and DA levels in AD as compared to CON when DOPA was adjusted were unexpected. Despite certain shared epidemiological, clinical, and neuropathologic features, evidence for dopaminergic pathology in AD has not been consistent and conclusive (60). A meta-analysis found lower levels of DA in patients with AD as compared with controls, however subanalysis showed that the difference was not significant in studies using brain tissues (61). Transgenic mice overexpressing a mutated human amyloid precursor protein (APP), a model of AD, displayed age-dependent loss of dopaminergic neurons in ventral tegmental area (VTA) but not SN (62). While not as great as in PD patients, loss of dopaminergic neurons has been reported in AD patients (63), especially loss of VTA volume appeared to correspond to hippocampal size and memory indices (64). No SN pathology was reported in the present study in all but one of the AD patients, who had mild SN neuron loss but not the lowest DA and DOPA values in the group. Except for a trend for reduced PTCA, we did not find changes in NM content or any of the eumelanin and pheomelanin markers in AD as compared with CON. Compared to PD, AD had lower 4-AHP/4-AHPEA, lower 4-AHP level as well as lower ratio of 4-AHP/PTCA. It is unclear whether AD shares common pathological mechanisms to PD, including early neurochemical abnormality in the SN but to a lesser extent or the NM changes are specific to PD.

Our study has limitations. First, the postmortem samples were race-, age-, and PMI-matched but not gender-matched. Given gender differences in both PD and AD (65), the results were gender adjusted, which were highly consistent with ANOVA results without controlling for gender. Due to the relatively small sample size, we did not stratify gender differences. Second, DOPA values from postmortem samples were total DOPA, which consisted of not only free DOPA but also Cys-DOPA, protein conjugated DOPA, and other forms of DOPA. Third, although our chemical methods for postmortem tissue assessments were well-validated, the values reported here were degradation markers for pheomelanin and eumelanin, not actual levels of pheomelanin and eumelanin. Lastly, although our previous studies did not identify differences in dopaminergic cell survival and microglial activation between gold suspension control and PBS, we cannot exclude in our αSyn mouse model possible particle effects from melanin suspension (56), especially, eumelanin is more insoluble than pheomelanin. In addition, our findings using synthetic DOPA pheomelanin and eumelanin may not model pheomelanin and eumelanin functions/structure in the human brain, which can be influenced by lipids, proteins, metals, and other components that are well identified components of human NM and NM-containing organelles (2, 7, 15, 66). Iron, for example, consists of 1% of NM by weight, which mostly binds to the eumelanic component. Iron is the main source of oxidative stress, especially when NM is released by dying neurons and interacts with the toxic species like H_2_O_2_ produced by activated microglia (7, 9). Nevertheless, our findings provide insights into the distinct roles of pheomelanin and eumelanin in PD pathophysiology and they form a foundation for further investigation on possible intervention to the dysregulated NM pathways in PD. With advances in the development of NM detection methods (67, 68), it may be possible and necessary to distinguish pheomelanin and eumelanin in the human brain as a biomarker for oxidative stress status, aging, and PD. The distinct functions of pheomelanin and eumelanin in the periphery have been extensively probed. High pheomelanin and related oxidative stress in redhaired people carrying loss-of-function variants of *MC1R* (melanocortin-1-receptor), which activates tyrosinase and facilitates eumelanin synthesis (69), have been associated with compromised drug and metal ion binding, abnormal skin sun damage, UV-independent skin aging as well as melanoma risk (24, 44, 70). We have reported the compromised nigrostriatal dopaminergic system in redhead mice and increased PD risk in redhaired people (30, 71). Further investigations should be warranted to better understand whether/how systemic pheomelanin might relate to pheomelanin in the NM of CNS and how it may affect dopaminergic neuron survival.

## 5. Conclusions

Dopaminergic neurons in the brain contains neuromelanin (NM) and are particularly vulnerable in Parkinson disease (PD). NM pigment consists of pheomelanin and eumelanin, which possess distinct chemical and biological characteristics. We found increased pheomelanin and reduced eumelanin in PD brain as compared to control subjects. In a mouse model of PD, pheomelanin exacerbated neurotoxicity while eumelanin attenuated dopamine deficit. Our study provides insights into the different roles of pheomelanin and eumelanin moieties in PD pathophysiology. It forms a foundation for further investigations on pheomelanin and eumelanin individually as biomarkers and therapeutic targets for PD.

## Data Availability

All data produced in the present work are contained in the manuscript.

## Abbreviations

AAV: Adeno-associated virus
AD: Alzheimer’s disease
3-AHP: 3-Amino-4-hydroxyphenylalanine
4-AHP: 4-Amino-3-hydroxyphenylalanine
3-AHPEA: 3-Amino-4-ydroxyphenylethylamine
4-AHPEA: 4-Amino-3-hydroxyphenylethylamine
AHPO: Alkaline hydrogen peroxide oxidation
BT: Benzothiazine
BZ: Benzothiazole
CON: Control
Cys: Cysteine
Cys-DA: Cysteinyldopamine
Cys-DOPA: Cysteinyldopa
DA: Dopamine
DOPA: L-3,4-dihydroxyphenylalanine
DOPAC: 3,4-dihydroxyphenylacetic acid
ECD: Electrochemical detection
HPLC: High performance liquid chromatography
HI: Hydroiodic acid
Iba1: Ionized calcium binding adapter molecule 1
NM: Neuromelanin
p-αSyn: Phosphorylated alpha-synuclein
PD: Parkinson’s disease
PDCA: Pyrrole-2,3-dicarboxylic acid
PMI: Postmortem interval
PTCA: Pyrrole-2,3,5-tricarboxylic acid
SN: Substantia nigra
SNpc: SN pars compacta
αSyn: alpha-Synuclein
TH: Tyrosine hydroxylase
TTCA: Thiazole-2,3,5-tricarboxylic acid
UVD: UV detection.

## Authors’ contributions

Conceptualization: WC, KW, SI, LZ, XC

Methodology: WC, KW, FAZ, KY, PS, SI, LZ, XC

Investigation: WC, KW, FAZ, KY, PS, LC, SI, LZ

Visualization: WC, KW, FAZ, KY, NM, PS, SI, LZ, XC

Funding acquisition: WC, KW, SI, LZ, XC

Project administration: XC

Supervision: SI, LZ, XC

Writing: original draft: KW, NM, XC

Writing: review & editing: WC, KW, FAZ, KY, PS, LC, SI, LZ, XC

## Data and materials availability

All data are available in the main text.

## Acknowledgements

We thank the research subjects who donated the tissues for this study and the Harvard Brain Tissue Resource Center/NIH NeuroBioBank for providing the samples. Statistical analysis was conducted in consultation with the Harvard Catalyst Biostatistics Consulting Program and Dr. Jian Wang at The University of Texas MD Anderson Cancer Center through the Aligning Science Across Parkinson’s (ASAP) initiative.

