## Supplemental Table 1 for "DOPA pheomelanin is increased in nigral neuromelanin of Parkinson’s disease and it exacerbates alpha-synuclein neurotoxicity"

**Supplementary materials**

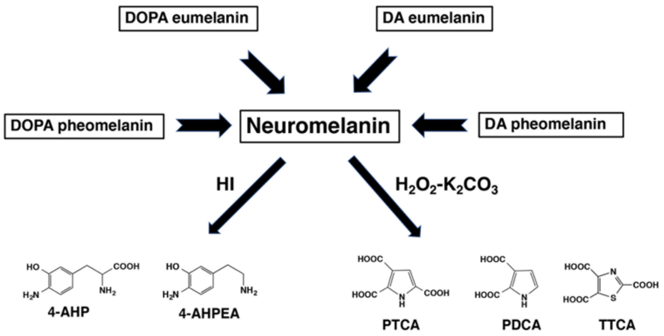


**Supplementary Figure 1: Chemical degradation of pheomelanin and eumelanin in the SN.** HI reduction produces 4-AHP and 4-AHPEA, markers for DOPA pheomelanin and DA pheomelanin, respectively. $H_2O_2$-$K_2CO_3$ oxidation (AHPO) produces PTCA, a marker for DOPA eumelanin, PDCA, a marker for DA eumelanin, and TTCA, a marker for DA pheomelanin.
